# ChatGPT Provides Inconsistent Risk-Stratification of Patients With Atraumatic Chest Pain

**DOI:** 10.1101/2023.11.29.23299214

**Authors:** Thomas F. Heston, Lawrence M. Lewis

**Affiliations:** Department of Medical Education and Clinical Sciences, Washington State University, Spokane, Washington, USA, and the Department of Family Medicine, University of Washington School of Medicine, Seattle, Washington, USA; Department of Emergency Medicine, Washington University, Saint Louis, Missouri, USA (ret.)

**Keywords:** artificial intelligence, large language models, ChatGPT, risk assessment, acute coronary syndrome

## Abstract

**BACKGROUND:** ChatGPT is a large language model with promising healthcare applications. However, its ability to analyze complex clinical data and provide consistent results is poorly known. This study evaluated ChatGPT-4’s risk stratification of simulated patients with acute nontraumatic chest pain compared to validated tools.

**METHODS:** Three datasets of simulated case studies were created: one based on the TIMI score variables, another on HEART score variables, and a third comprising 44 randomized variables related to non-traumatic chest pain presentations. ChatGPT independently scored each dataset five times. Its risk scores were compared to calculated TIMI and HEART scores. A model trained on 44 clinical variables was evaluated for consistency.

**RESULTS:** ChatGPT showed a high correlation with TIMI and HEART scores (r = 0.898 and 0.928, respectively), but the distribution of individual risk assessments was broad. ChatGPT gave a different risk 45-48% of the time for a fixed TIMI or HEART score. On the 44 variable model, a majority of the five ChatGPT models agreed on a diagnosis category only 56% of the time, and risk scores were poorly correlated (r = 0.605). ChatGPT assigned higher risk scores to males and African Americans.

**CONCLUSION:** While ChatGPT correlates closely with established risk stratification tools regarding mean scores, its inconsistency when presented with identical patient data on separate occasions raises concerns about its reliability. The findings suggest that while large language models like ChatGPT hold promise for healthcare applications, further refinement and customization are necessary, particularly in the clinical risk assessment of atraumatic chest pain patients.

## Introduction

The feasibility of using artificial intelligence (AI) to improve healthcare has only recently been made possible by revolutionary advances in neural network architecture. Neural networks were first applied to machine learning in the 1940s, starting with a single-neuron model (1). Computer neural networks gradually increased in complexity for decades, but only recently have advances in computer processing speeds coupled with the growth in the Internet resulted in programs able to communicate directly with humans using natural language (2). The transformer architecture developed in 2017 was a significant step forward in the language ability of neural networks by getting the programs to understand word context (3). Then, in 2018, the generative pre-trained transformer (GPT) model was able to generate coherent text (4).

Since the 2018 GPT-1 version, this model has been continuously improved to the point where an Internet-based chatbot, ChatGPT-4, can carry on natural conversations with humans in multiple languages, including spoken languages, archaic languages, and software coding languages. All iterations of these GPT large language models (LLMs) have “attention” mechanisms that allow the model to focus on specific input from which to “learn” its assigned task, thus making the neural network capable of learning from raw, unlabeled data. Previously, neural networks required labeled data from which to learn, requiring someone to view and interpret it before using it as input. While ChatGPT can carry on general conversations in multiple languages, its ability to reason and understand the language of medicine needs further evaluation. Multiple studies show that ChatGPT does well on single medical task commands such as answering multiple-choice examination questions such as the United States Medical Licensing Exam (5,6) and medical specialty exams (6). However, ChatGPT needs help with logical questions (7) and tends to fabricate answers (8). It can generate convincing, misleading text containing glaring errors (9).

One area in medicine that has extensively been at the forefront of AI usage is diagnostic imaging. Recent studies looking at the performance of AI to identify a specific or small number of pathologic conditions have shown AI to be as accurate as trained physicians for interpreting specific radiographic procedures (10), although not as accurate as the practice of double-reading (11). Clinical decision-making and risk stratification present a different level of complexity, requiring input from the history, examination, laboratory results, and imaging studies. Nonetheless, several studies have found AI (in particular machine learning) performed as well or better than the standard risk-stratification tools currently in use for conditions such as transcatheter aortic valve implantation (12), surgical risk (13), and cardiovascular risk (14–17) Most of these studies used machine learning to incorporate and integrate labeled data from clinical registries, including outcome data, into their prediction model.

Chest pain is a common chief complaint in the emergency department, which can be associated with severe medical conditions but usually has a benign cause. Physician caution has led to many patients being admitted to rule out acute coronary syndrome that do not have heart disease. This over-use of resources led to efforts to develop accurate risk-stratification protocols to better identify low-risk patients. One such protocol is the TIMI score, a seven-item screening tool designed to identify people at low risk for a major adverse outcome at 14 days (18). Another protocol is the HEART score. This five-item screening tool predicts the six-week risk of major adverse cardiac events (19).

In this study, we looked at the ability of ChatGPT-4 to evaluate clinical scenarios involving a series of simulated patients with acute nontraumatic chest pain. Its risk stratification performance was compared to TIMI and HEART scores. Then, to further investigate the behavior of ChatGPT-4, it was presented with more complex simulated patients to identify which variables it determined to be of primary importance for stratifying risk and to assess if its responses were consistent when presented with identical data on different occasions.

## Methods

### Data generation

Case studies were randomly simulated using a Python software program. All cases were computer-generated; no actual patient data was studied. There were three datasets of simulated case studies generated.

The first dataset contained the seven TIMI score variables for unstable angina or non-ST elevation myocardial infarction (18). To facilitate interaction with ChatGPT, the variables were coded in binary fashion with 0=no and 1=yes as follows: age >= 65 (yes/no); >= 3 coronary artery disease (CAD) risk factors (yes/no); known CAD (yes/no); aspirin use in the past seven days (yes/no); two or more episodes of severe angina within the past 24 hours (yes/no); EKG ST changes of 0.5 mm or more (yes/no); and positive cardiac marker (yes/no).

The second dataset of simulated case studies consisted of the five HEART score variables for major cardiac events (19) as follows: history (slightly suspicious, moderately suspicious, or highly suspicious); EKG (normal, non-specific repolarization disturbance, or significant ST deviation); age (<45, 45-64, or over 64); risk factors (no known risk factors, one or two risk factors, three or more risk factors); and initial troponin (normal, one to three times normal limit, or over three times the normal limit).

The third dataset of simulated cases consisted of forty-four randomized variables related to the acute presentation of non-traumatic chest pain. These variables were selected to represent common conditions in acute nontraumatic chest pain patients. This dataset of simulated patients only included variables related to the history and physical. It did not include test results. These variables were age (40 to 90), duration of pain in minutes, pain level of severity (1 to 10), gender (M/F), race (African American or non-African American); and the following binary variables randomly coded as 0=no, 1=yes: substernal chest pain, heavy pain, burning type of pain, pain precipitated by exertion or stress, pain relieved by rest or nitroglycerin, pain worse with lying down, pain worse with deep breath, currently on aspirin, currently on a blood pressure medication, currently on a nonsteroidal anti-inflammatory medication, currently on a statin medication, currently on insulin, uses cocaine, moderate to heavy alcohol use, current smoker, history of hypertension, previous myocardial infarction, history of diagnosed coronary artery disease, history of diabetes, history of stroke, currently experiencing nausea, currently experiencing dyspnea, currently experiencing palpitations, currently experiencing dizziness, currently married, family history of coronary artery disease, currently hypotensive on exam, currently hypertensive on exam, currently bradycardic on exam, currently tachycardic on exam, currently febrile on exam, currently tachypneic on exam, currently hypoxic on exam, weak pulse on exam, irregular heart rhythm on exam, abnormal lung sounds on auscultation, pain reproducible on palpation, murmur present on cardiac auscultation, and edema present on exam.

### ChatGPT analysis

The three datasets were processed individually. First, the dataset was uploaded to ChatGPT-4 (September 25, 2023 Version) Advanced Data Analysis. A standardized set of prompts was utilized to interact with ChatGPT. First, ChatGPT was prompted to assign each case a risk score for acute coronary syndrome. The risk score for the first dataset was set to a scale from 0 to 7 (corresponding to the TIMI scale); for the second dataset, it was scaled from 0 to 10 (corresponding to the HEART scale); and for the final dataset, the scale was set from 0 to 100. Finally, ChatGPT was instructed to show the weighting it applied to each variable when calculating its risk score. For the third dataset, ChatGPT was instructed to give a weight to each of the 44 variables; a weight of zero was explicitly allowed.

For the third set of simulated cases, ChatGPT was also prompted to state the first test it would order in the emergency department for each case. This was an open-ended prompt in which no specific guidelines or limitations were given.

Each dataset was presented to ChatGPT five times, with instructions to assign a risk score to each case and show the weight it gave to each variable. Datasets were not changed, meaning the same data was presented to ChatGPT on five occasions in a new chat session. This created five models for each of the three datasets, creating a total of 15 models. A new chat session was initiated each time when creating a new model. Each of the five models for the third dataset also included a recommendation for the first test to order in the emergency department for each case. These models were named 1.1, 1.2, 1.3, 1.4, and 1.5 for the first dataset. Similar naming was used for the five models for the second dataset and the five models for the third dataset. Finally, only after the ChatGPT models had assigned risk scores was the TIMI score calculated for each simulated patient in the first dataset and the HEART score calculated for each simulated patient in the second dataset.

After this process, the first dataset consisted of the seven TIMI variables, the TIMI score, and five risk scores (one score for each of the five ChatGPT models generated). The second dataset consisted of the five HEART variables, the HEART score, and five risk scores from each of the five models developed. The third dataset consisted of forty-four variables, five risk scores (one from each model), and five “first test” recommendations (one for each ChatGPT model). The weights assigned to each variable by each model were recorded.

### Statistical analysis

The three datasets were evaluated using IBM SPSS Statistics (Version 29). Excel was used for calculating averages of weighting scores. Pearson’s correlation coefficient and R-squared were calculated to compare the risk scores of each model. The recommended first test was stratified by gender and race, and a test of proportions was performed to analyze any differences. The Kruskal-Wallis test was used to determine if the various models created by ChatGPT for each dataset were statistically different. The SAMPL guidelines were used for the design and analysis of this study (20).

### Data availability

The Python code, ChatGPT prompts, the three final datasets, and the SPSS output files are all open-access and deposited in the Zenodo repository (21).

## Results

ChatGPT had difficulty assigning risk scores corresponding to the TIMI and HEART scales, frequently giving condensed or expanded scales. Prompting was therefore changed to have ChatGPT provide a risk score from 0 to 100 inclusive, and in this situation, it did not have any difficulty. These risk scores were then adjusted to match the TIMI and HEART scales.

For the third dataset, which included forty-four variables from a simulated patient’s history and physical, ChatGPT frequently took shortcuts in assigning weights, responding that “for convenience” or “for brevity,” it was setting a default weight to multiple variables. For this dataset, prompting was modified to force ChatGPT to independently evaluate every variable and give a weight to come up with a risk score and its recommendation for the first test to order in the emergency department. Negative weights or weights of zero were explicitly allowed. The third dataset presented to ChatGPT for final analysis had all variables ranging from zero to one except for age (range 40 to 90 years), duration of pain (1 to 40 minutes), and severity of pain (1 to 10).

### TIMI dataset

There were 10,000 simulated cases generated for the TIMI dataset. The TIMI scores were normally distributed (Figure 1). Correlation coefficients between the TIMI scores and the five ChatGPT models were high and, in all cases, statistically significant, with a p-value of < 0.001 (Table 1). Overall, the correlation between TIMI and ChatGPT was 0.898 (p < 0.001).

**Figure 1.**
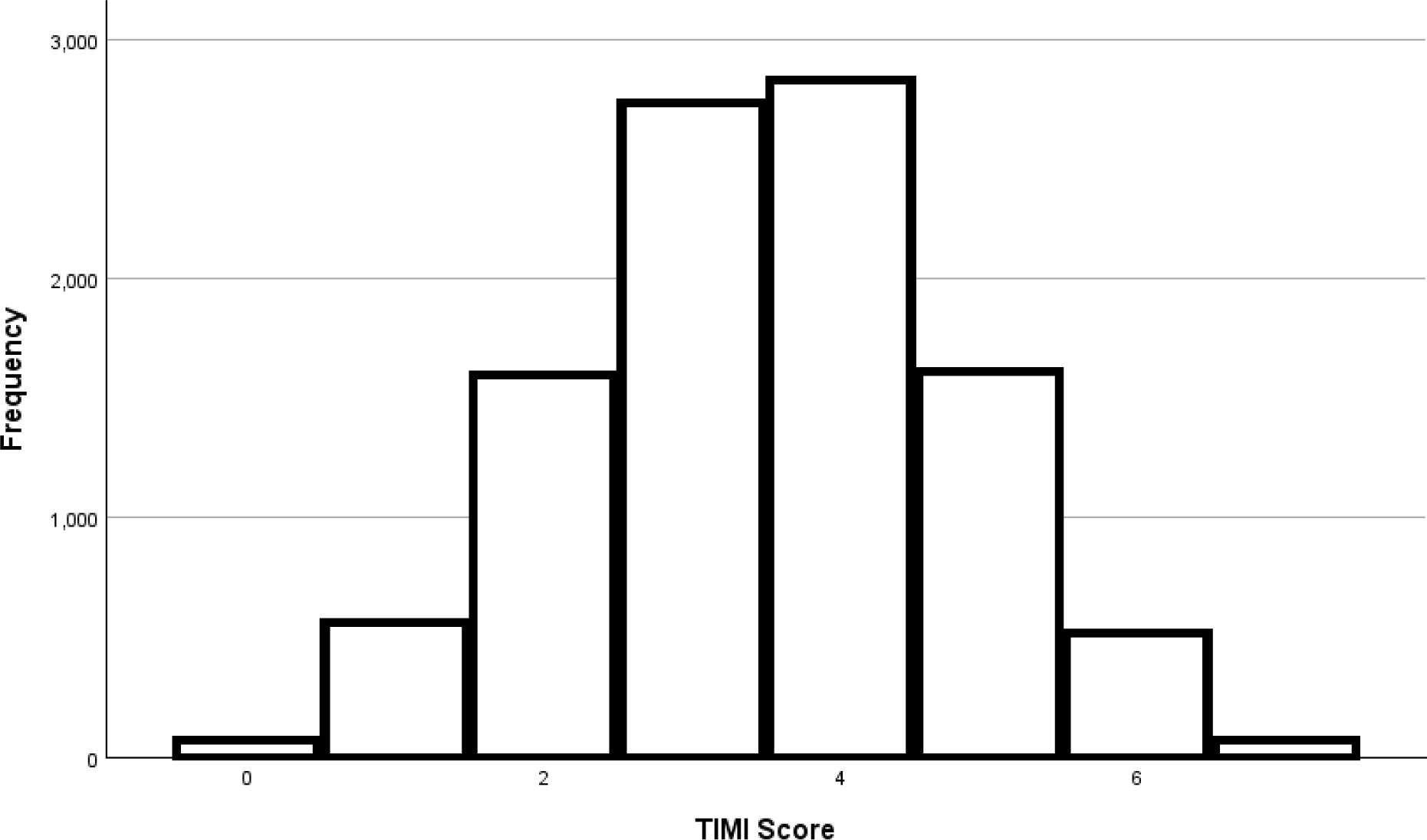
Histogram of TIMI scores in the first dataset The TIMI dataset was normally distributed.

**Table 1.** TIMI Dataset Correlation Matrix Pearson correlation coefficients were 0.815 or greater in all cases. All correlation coefficients were statistically significant with p < 0.001.

| Model | TIMI | Model 1.1 | Model 2.1 | Model 3.1 | Model 4.1 |
| --- | --- | --- | --- | --- | --- |
| Model 1.1 | 0.930 | - | - | - | - |
| Model 1.2 | 0.900 | 0.921 | - | - | - |
| Model 1.3 | 0.899 | 0.916 | 0.949 | - | - |
| Model 1.4 | 0.851 | 0.931 | 0.862 | 0.877 | - |
| Model 1.5 | 0.908 | 0.894 | 0.936 | 0.958 | 0.815 |

Although the correlations were uniformly high, the data showed a broad distribution when comparing ChatGPT scores with TIMI scores. For TIMI scores of zero and seven, as expected, there was perfect agreement with the ChatGPT models since only seven variables were evaluated by these models; for a TIMI score of zero, all variables would be negative, so in all cases, the weighting applied by ChatGPT to each variable would sum to zero. Similarly, all variables would be positive for a TIMI score of seven, so the weighting applied by ChatGPT would add to its maximum. The mean (standard error) for TIMI was 3.50 (0.013), and for ChatGPT was 3.55 (0.015), which was a statistically significant difference by paired t-test (p = < 0.001). The ChatGPT models gave a different score than TIMI 45% of the time. ChatGPT gave three to four different scores for a fixed TIMI score from one to six (Figure 2).

**Figure 2.**
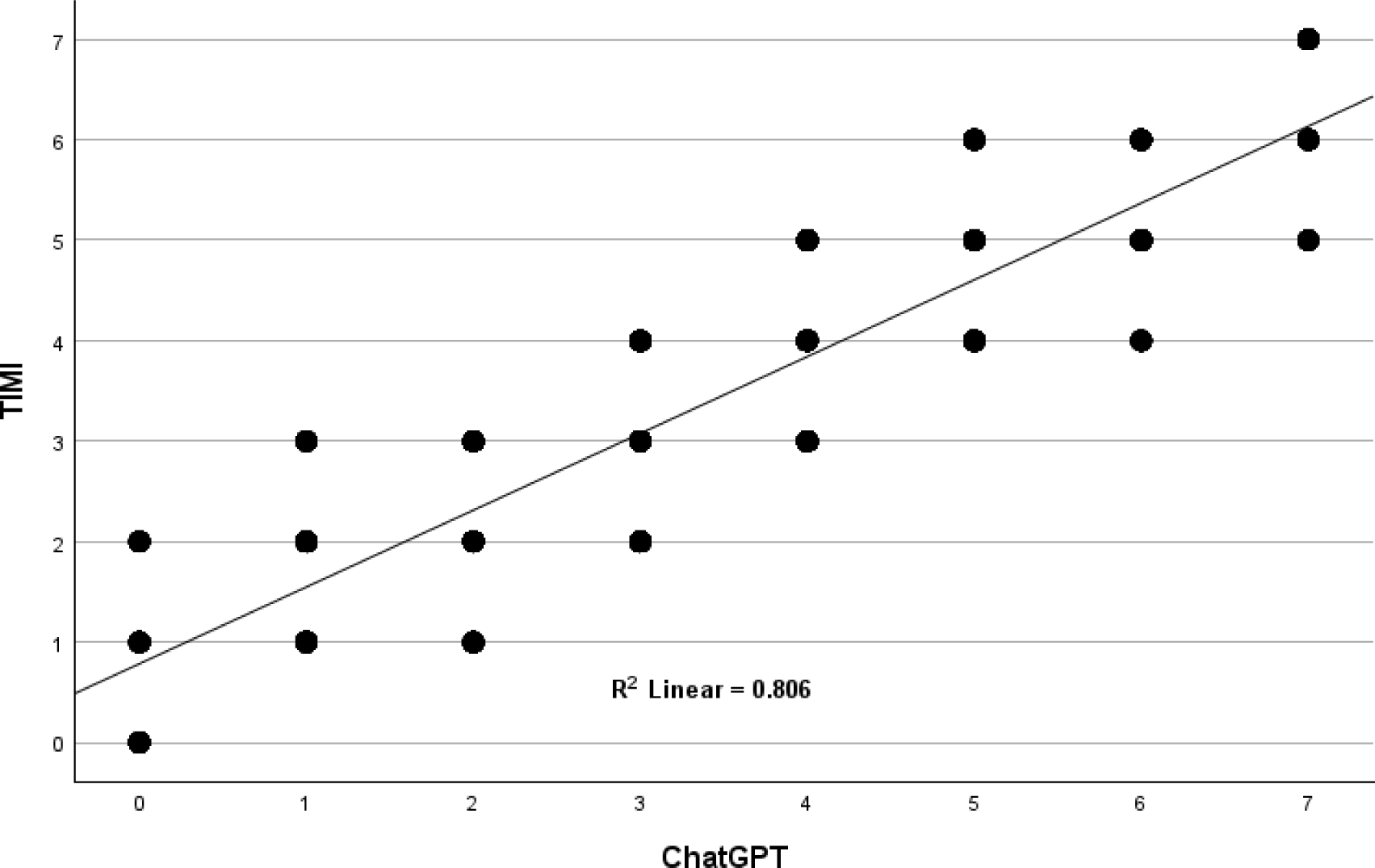
Comparison of TIMI with ChatGPT The correlation between TIMI and ChatGPT was high, with an R-squared of 0.806; however, the data distribution was broad.

The weights assigned to each variable by the five ChatGPT models were statistically similar (p=0.406 by Kruskal-Wallis). However, there were different weights assigned by the various models for each variable. The specific weights assigned to each TIMI variable by each of the five ChatGPT models generated are listed in Table 3.

**Table 3.**
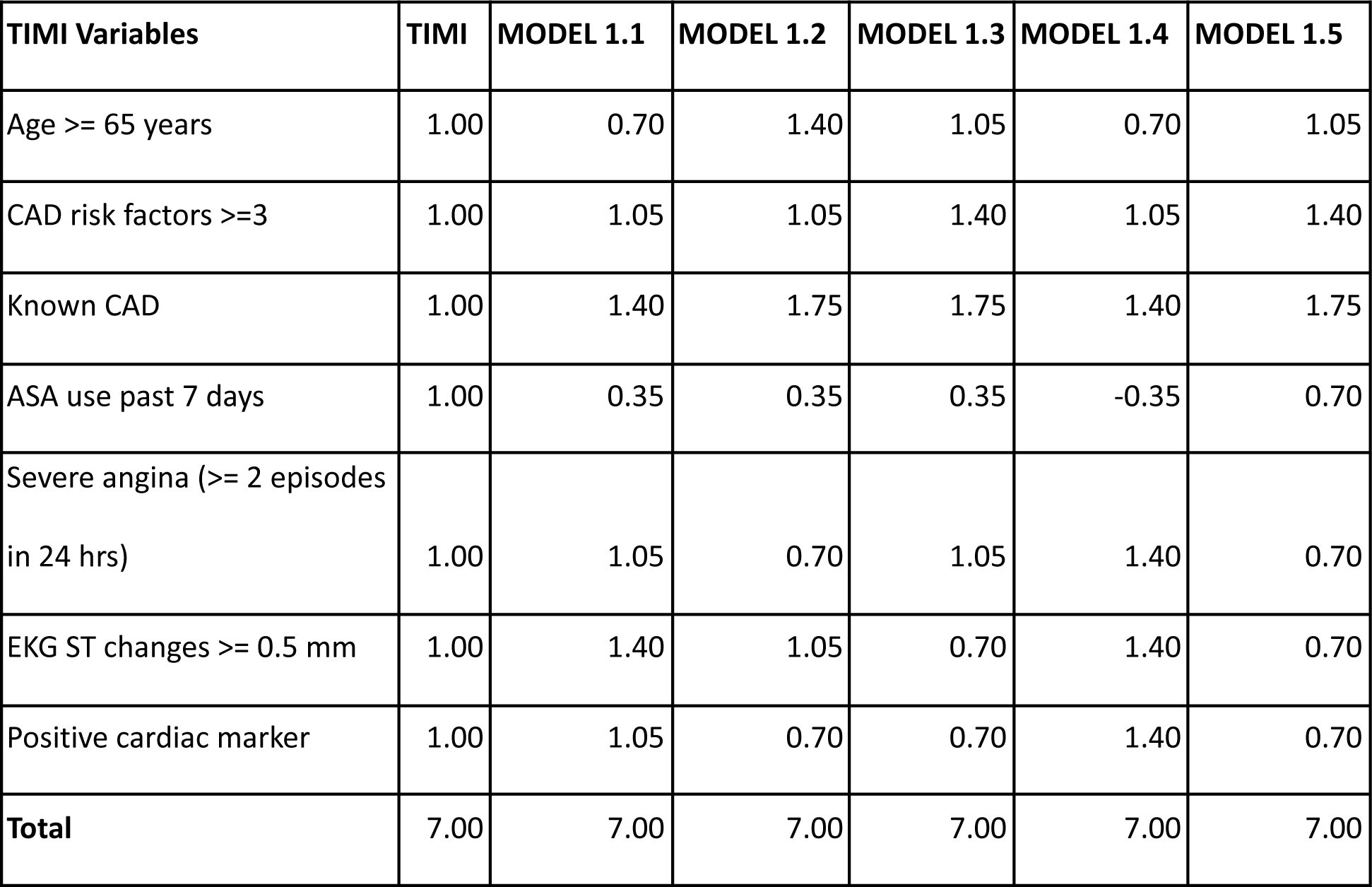
Weights assigned to the TIMI variables by the five ChatGPT models. CAD = coronary artery disease, ASA = aspirin, a positive cardiac marker refers to an elevated CKMB or troponin level.

### HEART dataset

There were 10,000 simulated cases generated for the HEART dataset. The HEART scores were normally distributed (Figure 3). Correlation coefficients between the HEART scores and the five ChatGPT models were high and, in all cases, statistically significant, with a p-value of < 0.001 (Table 4). The correlation between HEART and ChatGPT overall was 0.928 (p < 0.001).

**Figure 3.**
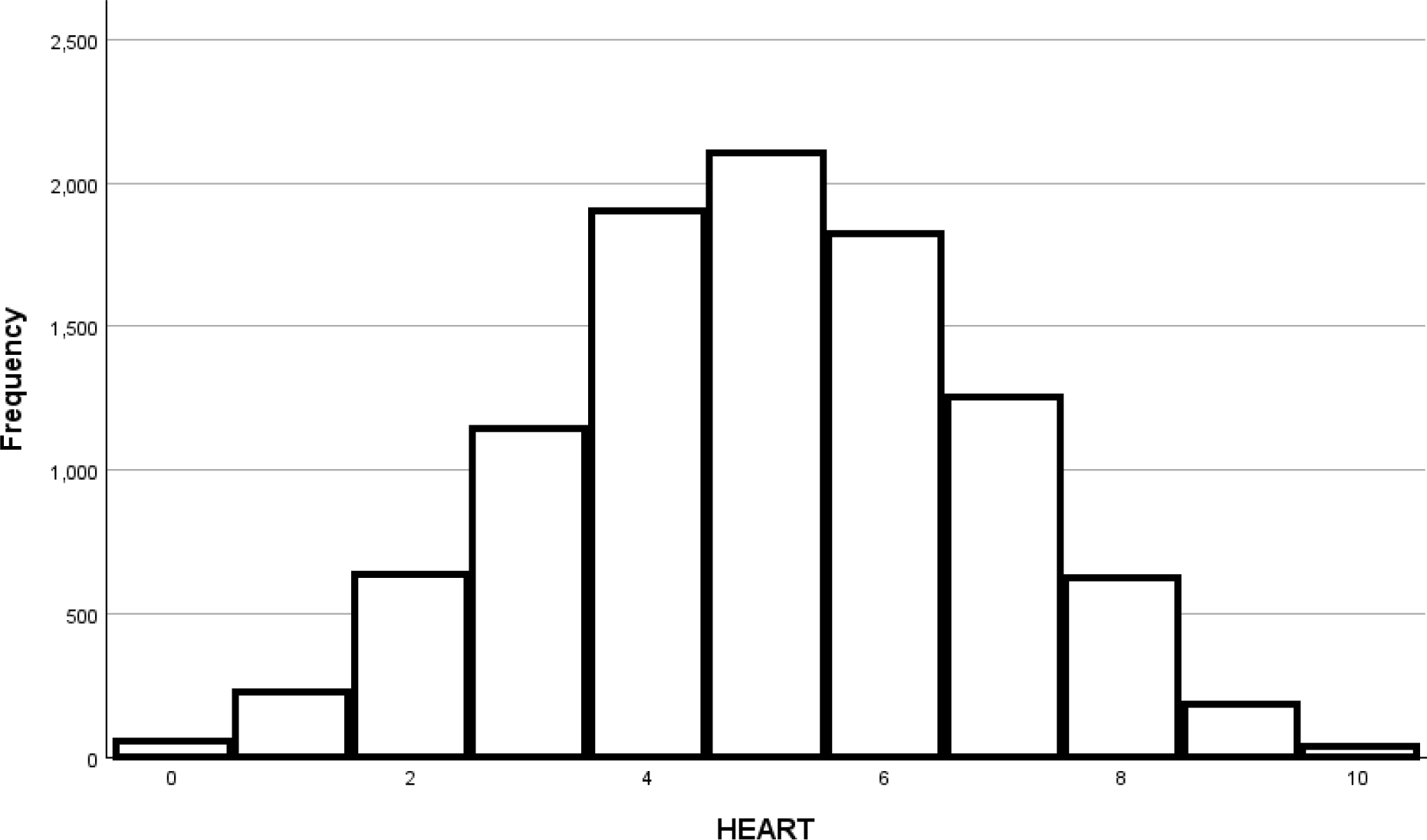
Histogram of HEART scores in the second dataset The HEART dataset was normally distributed.

**Table 4.** HEART dataset correlation matrix Pearson correlation coefficients were 0.865 and greater in all cases. All correlation coefficients were statistically significant with p < 0.001.

| Model | HEART | Model 2.1 | Model 2.2 | Model 2.3 | Model 2.4 |
| --- | --- | --- | --- | --- | --- |
| Model 2.1 | 0.912 | - | - | - | - |
| Model 2.2 | 0.936 | 0.961 | - | - | - |
| Model 2.3 | 0.956 | 0.874 | 0.865 | - | - |
| Model 2.4 | 0.946 | 0.950 | 0.951 | 0.879 | - |
| Model 2.5 | 0.953 | 0.979 | 0.981 | 0.912 | 0.947 |

Similar to the TIMI analysis, correlations between the ChatGPT models and the HEART score were uniformly high, but the ChatGPT data showed a much broader distribution when compared with HEART scores. For HEART scores of zero and ten, as expected, there was perfect agreement with the ChatGPT models since only ten variables were evaluated by these models. The mean (standard error) HEART score was 4.99 (0.018), and for ChatGPT was 4.92 (0.020), which was a statistically significant difference by paired t-test (p < 0.001). However, of more concern, the ChatGPT models gave a broad range of scores for HEART scores from one to nine (Figure 4). Overall, the ChatGPT score differed from the HEART score 48% of the time.

**Figure 4.**
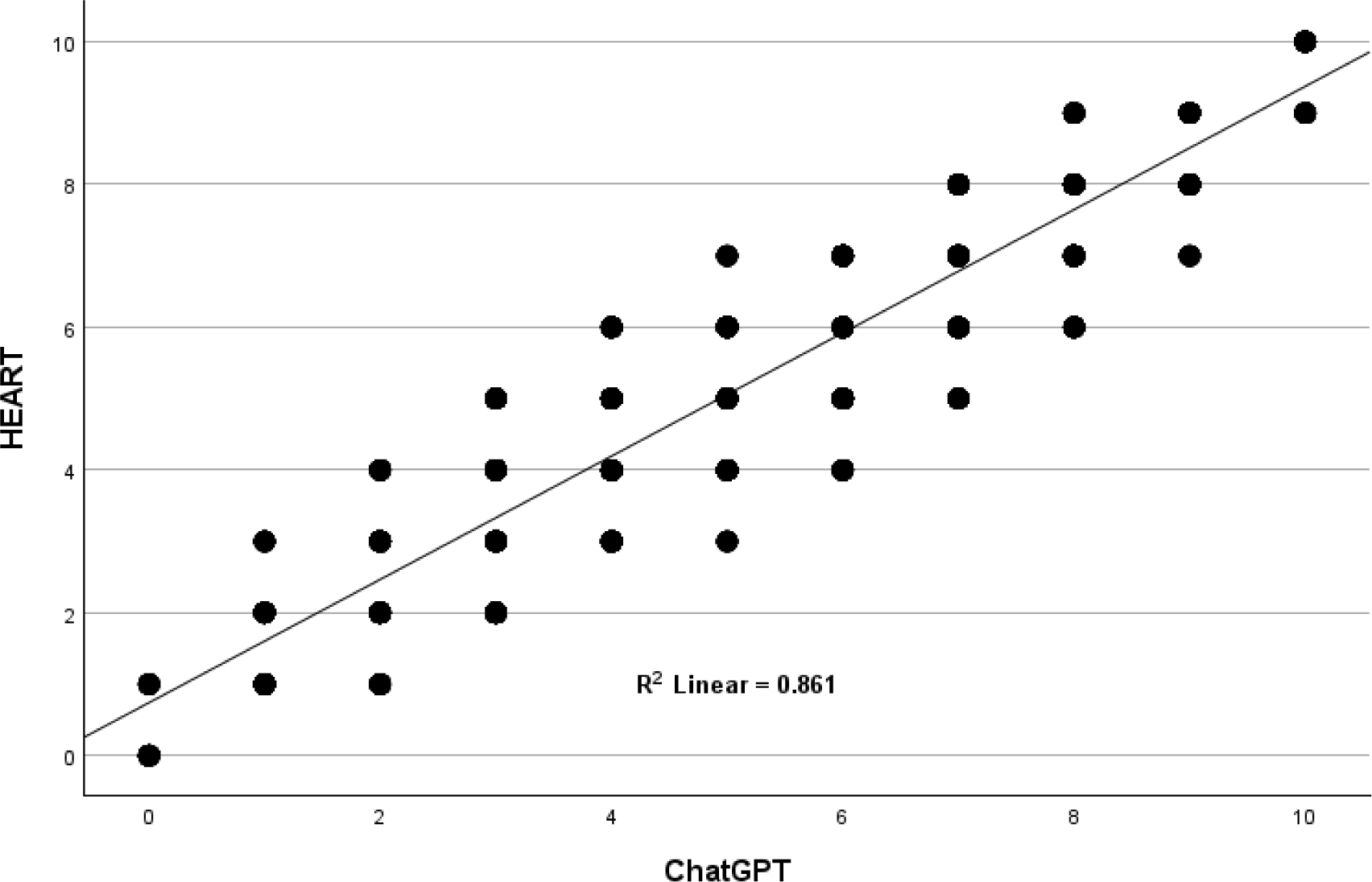
Comparison of HEART with ChatGPT The correlation between ChatGPT and HEART scores was high, with an R-squared of 0.861; however, the data distribution was broad.

When grouped by risk, the HEART variables encompass three categories: low, moderate, and high. When using a low-risk HEART score as the gold standard, ChatGPT had a sensitivity of 88%, a specificity of 93%, a positive predictive value of 76%, and a negative predictive value of 97% (Table 5). Overall concordance between HEART and ChatGPT across all 10,000 simulated patients, all three risk categories, and all five models ranged from 39% to 89% (Table 6).

**Table 5.** Performance of ChatGPT in Predicting a Low-Risk HEART score. the predictive value of a low-risk ChatGPT score indicating a low-risk HEART score was 76%.

| Scoring System | HEART Low Risk | HEART Not Low Risk | Total |
| --- | --- | --- | --- |
| ChatGPT Low Risk | 1824 | 581 | 2405 |
| ChatGPT Not Low Risk | 238 | 7357 | 7595 |
| Total | 2062 | 7938 | 10000 |

**Table 6.** Overall concordance between HEART and ChatGPT. Risk Categories concordance of HEART and ChatGPT assigned risk scores ranged from 39% to 89%.

| Risk Category | Risk | Concordance |
| --- | --- | --- |
| **Low** | 0.9% - 1.7% | **75%** |
| **Moderate** | 12% - 16.6% | **89%** |
| **High** | 50% - 65% | **39%** |

The weights assigned to each HEART variable by the five ChatGPT models were statistically similar (p=0.277 by Kruskal-Wallis). However, the various models set notably different weights for each variable. (Table 7).

**Table 7.**
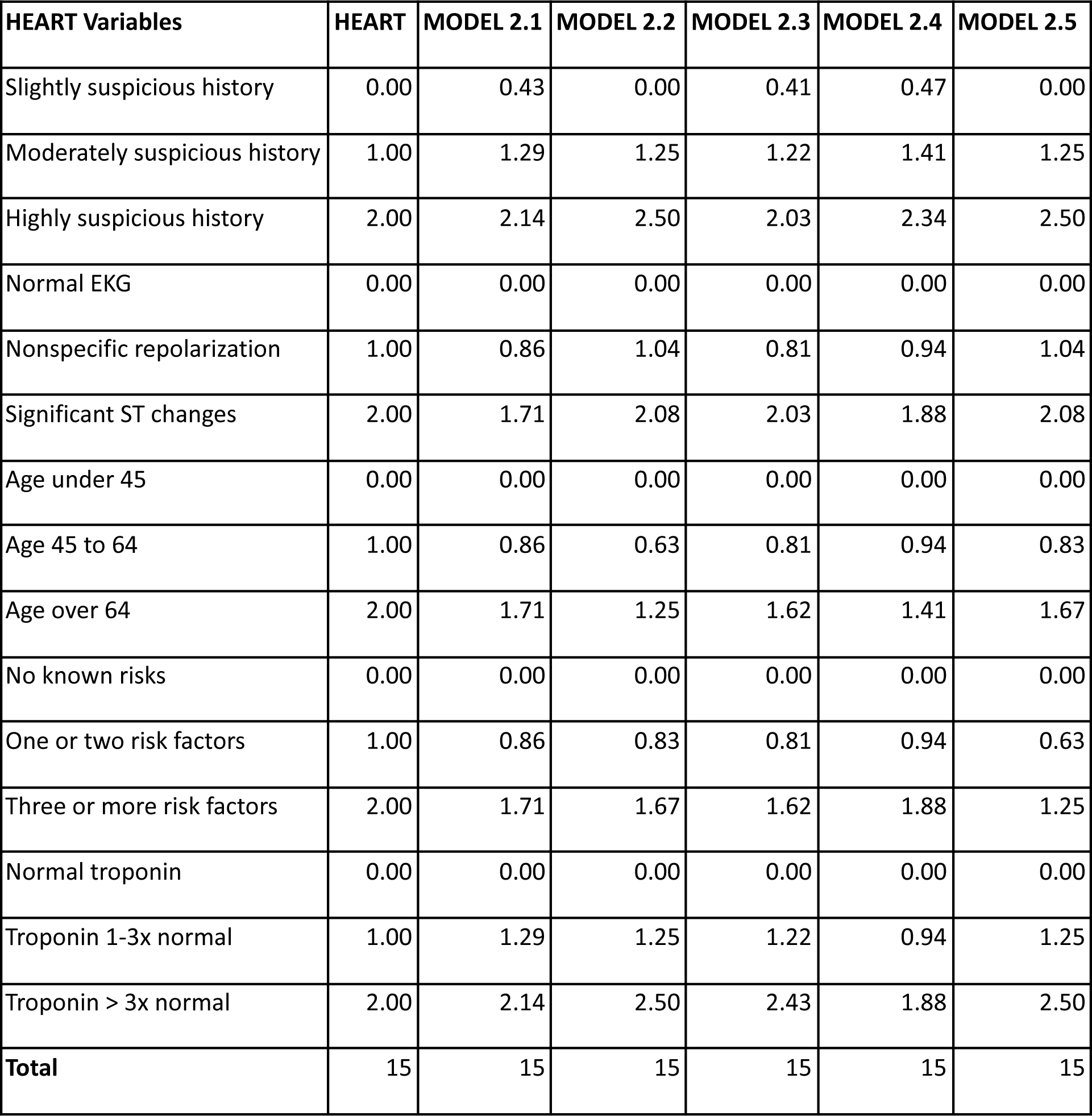
Weights assigned to the HEART variables by the five ChatGPT models. Although not statistically different, the five ChatGPT models assigned different weights to each variable.

### History and physical-only dataset

The risk scores were normally distributed, although slightly skewed left (Figure 5).

**Figure 5.**
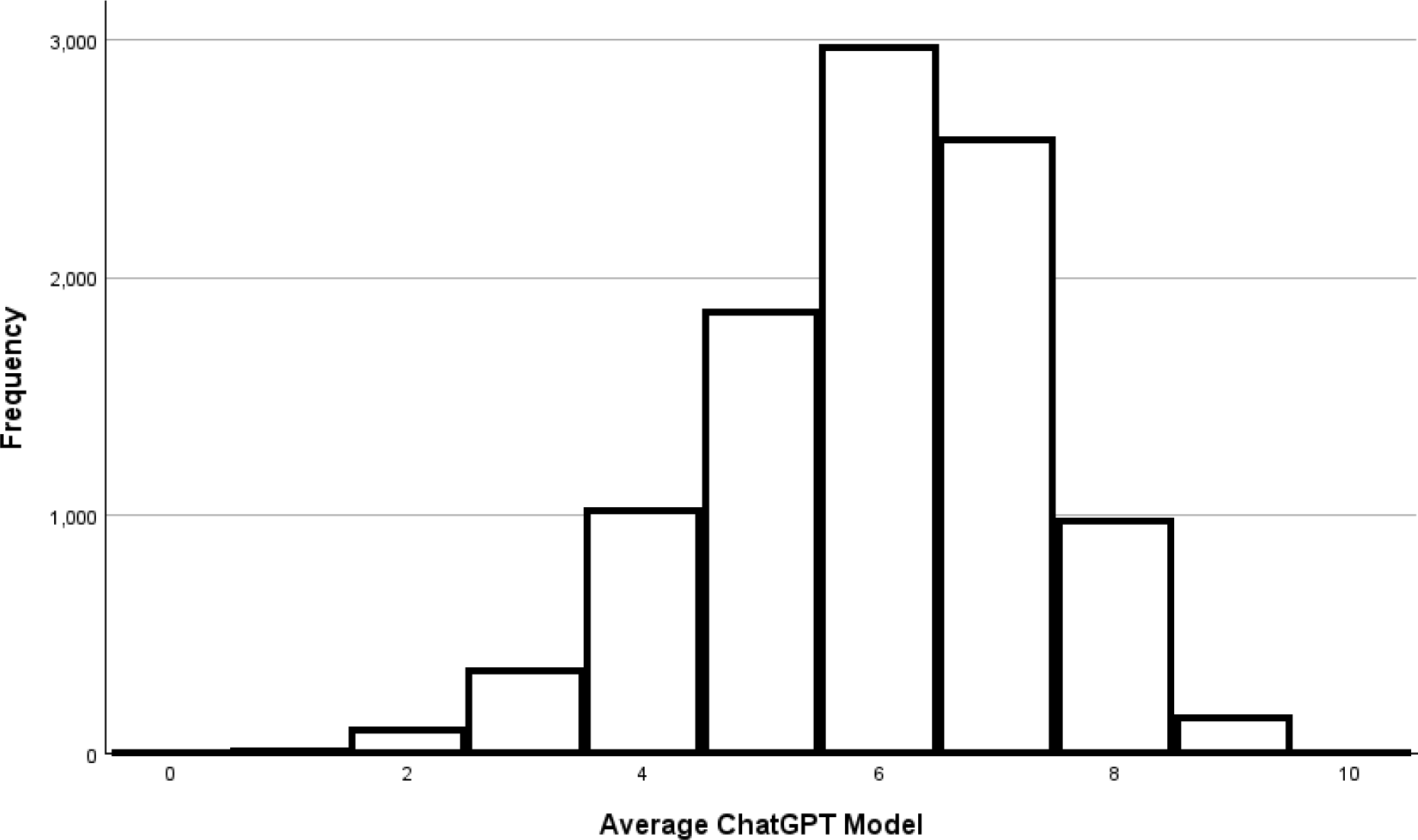
Risk scores assigned by ChatGPT to the history and physical dataset Risk scores assigned by ChatGPT to the history and physical-only dataset.

Given that there is no gold standard to compare the models built upon the history and physical-only dataset, the average score of the five models was utilized as a surrogate gold standard. The correlations with the average were all 0.808 or higher (Table 8).

**Table 8.**
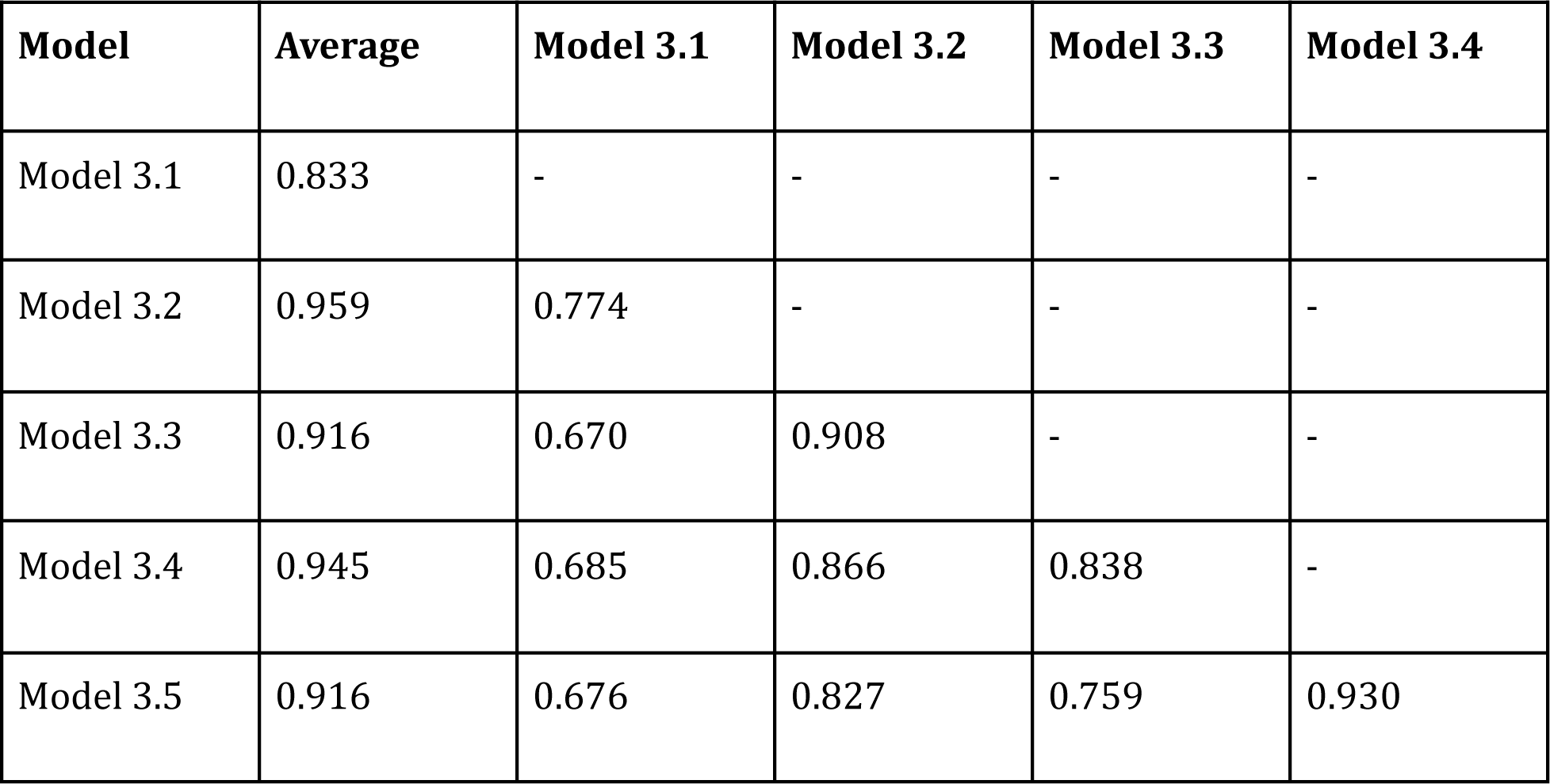
Correlation matrix for the history and physical-only dataset. Pearson correlation coefficients were 0.670 and greater in all cases. All correlation coefficients were statistically significant with p < 0.001.

A scatterplot comparing the individual model scores with the average score was generated to understand the distribution of risk scores across all five models (Figure 6). Note that while the individual correlations with the average risk score were high when the models were combined, there was a poor overall correlation (r = 0.605, R-squared = 0.366). For example, for an average score of four, the scores of the individual models ranged from two to nine.

**Figure 6.**
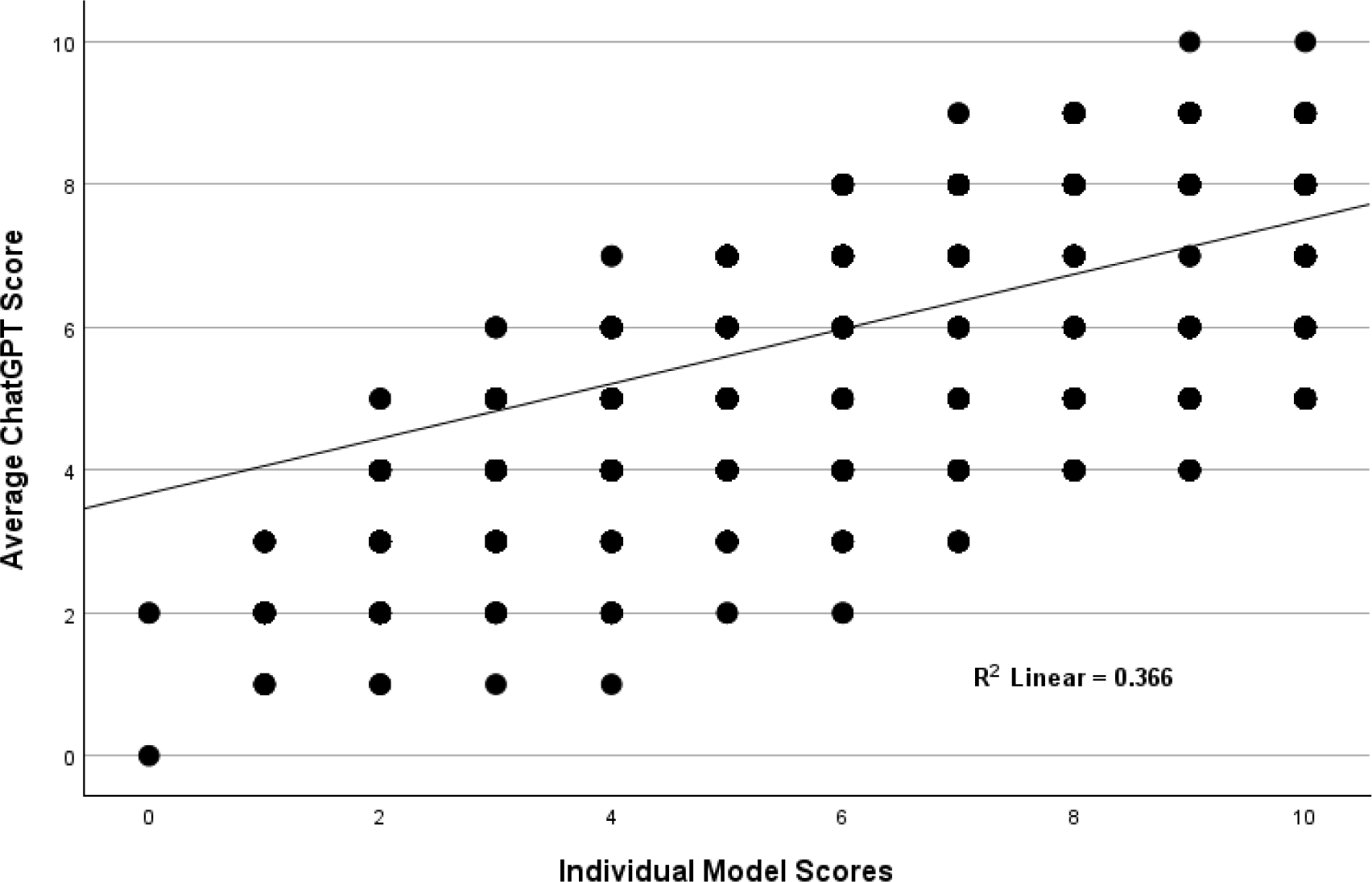
Individual model scores compared to average scores for the history and physical-only dataset

Age was given the highest weight by the ChatGPT models on average, contributing 8% to the overall risk score. The Diamond and Forrester criteria for chest pain were also assigned high weights on average (22). Pain precipitated by exertion or stress contributed 5.5%, pain relieved by rest or nitroglycerin contributed 5.1%, and a substernal location of pain contributed 5%. Pain reproducible on palpation and burning pain were protective factors, decreasing the cardiac risk scores (Table 9).

**Table 9.** Weights assigned by the history and physical dataset.

| **VARIABLE** | **Model 1** | **Model 2** | **Model 3** | **Model 4** | **Model 5** | **Average** |
| --- | --- | --- | --- | --- | --- | --- |
| Age | 0.093 | 0.063 | 0.111 | 0.060 | 0.071 | 0.080 |
| Pain Precipitated by Exertion or Stress | 0.037 | 0.057 | 0.089 | 0.048 | 0.043 | 0.055 |
| Pain Relieved by Rest or NTG | 0.037 | 0.057 | 0.078 | 0.042 | 0.043 | 0.051 |
| Heavy Pain | 0.037 | 0.044 | 0.078 | 0.042 | 0.050 | 0.050 |
| Substernal Chest Pain | 0.047 | 0.051 | 0.089 | 0.006 | 0.057 | 0.050 |
| Duration of Pain in Minutes | 0.023 | 0.032 | 0.067 | 0.054 | 0.035 | 0.042 |
| Pain Level of Severity | 0.047 | 0.025 | 0.067 | 0.036 | 0.028 | 0.041 |
| Male | 0.009 | 0.032 | 0.056 | 0.030 | 0.035 | 0.032 |
| Weak Pulse on Exam | 0.023 | 0.019 | 0.011 | 0.036 | 0.043 | 0.026 |
| Irregular Heart Rhythm on Exam | 0.023 | 0.019 | 0.011 | 0.024 | 0.050 | 0.025 |
| Uses Cocaine | 0.014 | 0.019 | 0.011 | 0.048 | 0.021 | 0.023 |
| Currently Tachycardic on Exam | 0.023 | 0.019 | 0.011 | 0.024 | 0.035 | 0.023 |
| History of Diagnosed Coronary Artery Disease | 0.019 | 0.019 | 0.011 | 0.042 | 0.021 | 0.022 |
| Previous Myocardial Infarction | 0.014 | 0.019 | 0.011 | 0.042 | 0.021 | 0.022 |
| Currently Hypoxic on Exam | 0.023 | 0.019 | 0.011 | 0.024 | 0.028 | 0.021 |
| Currently on Insulin | 0.023 | 0.019 | 0.011 | 0.030 | 0.021 | 0.021 |
| History of Hypertension | 0.019 | 0.019 | 0.011 | 0.024 | 0.028 | 0.020 |
| History of Diabetes | 0.019 | 0.019 | 0.011 | 0.030 | 0.021 | 0.020 |
| Edema Present on Exam | 0.023 | 0.019 | 0.011 | 0.018 | 0.028 | 0.020 |
| Currently Tachypneic on Exam | 0.023 | 0.019 | 0.011 | 0.018 | 0.028 | 0.020 |
| Currently Hypotensive on Exam | 0.023 | 0.019 | 0.011 | 0.024 | 0.021 | 0.020 |
| Currently Experiencing Dyspnea | 0.023 | 0.019 | 0.011 | 0.024 | 0.021 | 0.020 |
| Currently on a Statin Medication | 0.023 | 0.019 | 0.011 | 0.024 | 0.021 | 0.020 |
| Currently on Aspirin | 0.023 | 0.019 | 0.011 | 0.024 | 0.021 | 0.020 |
| Current Smoker | 0.014 | 0.019 | 0.011 | 0.030 | 0.021 | 0.019 |
| Family History of CAD | 0.019 | 0.019 | 0.011 | 0.024 | 0.021 | 0.019 |
| Currently Hypertensive on Exam | 0.023 | 0.019 | 0.011 | 0.018 | 0.021 | 0.019 |
| Currently Bradycardic on Exam | 0.023 | 0.019 | 0.011 | 0.018 | 0.021 | 0.019 |
| Currently Experiencing Dizziness | 0.023 | 0.019 | 0.011 | 0.018 | 0.021 | 0.019 |
| Currently Experiencing Nausea | 0.023 | 0.019 | 0.011 | 0.018 | 0.021 | 0.019 |
| Currently Experiencing Palpitations | 0.023 | 0.019 | 0.011 | 0.018 | 0.021 | 0.019 |
| Currently on a BP Medication | 0.023 | 0.019 | 0.011 | 0.018 | 0.021 | 0.019 |
| History of Stroke | 0.019 | 0.019 | 0.011 | 0.018 | 0.021 | 0.018 |
| Currently on an NSAID Medication | 0.023 | 0.019 | 0.011 | 0.012 | 0.021 | 0.017 |
| Currently Married | 0.023 | 0.019 | 0.011 | 0.006 | 0.021 | 0.016 |
| Murmur Present on Cardiac Auscultation | 0.014 | 0.019 | 0.011 | 0.012 | 0.021 | 0.015 |
| Moderate to Heavy Alcohol Use | 0.014 | 0.019 | 0.011 | 0.012 | 0.021 | 0.015 |
| Pain Worse With Deep Breath | 0.014 | 0.019 | 0.011 | 0.012 | 0.021 | 0.015 |
| Pain Worse With Lying Down | 0.014 | 0.019 | 0.011 | 0.012 | 0.021 | 0.015 |
| African American | 0.005 | 0.013 | 0.022 | 0.012 | 0.014 | 0.013 |
| Currently Febrile on Exam | 0.023 | 0.019 | 0.011 | 0.012 | -0.014 | 0.010 |
| Abnormal Lung Sounds on Auscultation | 0.014 | 0.019 | 0.011 | 0.012 | -0.021 | 0.007 |
| Pain Reproducible on Palpation | 0.014 | 0.019 | 0.011 | -0.030 | -0.057 | -0.009 |
| Burning Type of Pain | -0.023 | -0.019 | -0.033 | -0.030 | -0.035 | -0.028 |
| **Total** | **1.000** | **1.000** | **1.000** | **1.000** | **1.000** | **1.000** |

### Diagnoses and recommendations for initial test

ChatGPT was prompted to give the most likely diagnosis and the best initial test to order in the emergency department for the third dataset consisting of the history and physical variables. A wide range of diagnoses was given with slightly different wording by each of the models, e.g., one model would say “acute coronary syndrome,” and another model would say “ACS.” To simplify the data for analysis, the diagnoses were categorized as cardiovascular, pulmonary, gastrointestinal, musculoskeletal, or unknown.

For these categorized diagnoses, at least two models agreed on the initial diagnosis category over 99% of the time; three or more models agreed on the best diagnosis 56% of the time; four or more models agreed 22% of the time; and all five models agreed on the initial diagnosis category 5% of the time (Figure 7).

**Figure 7.**
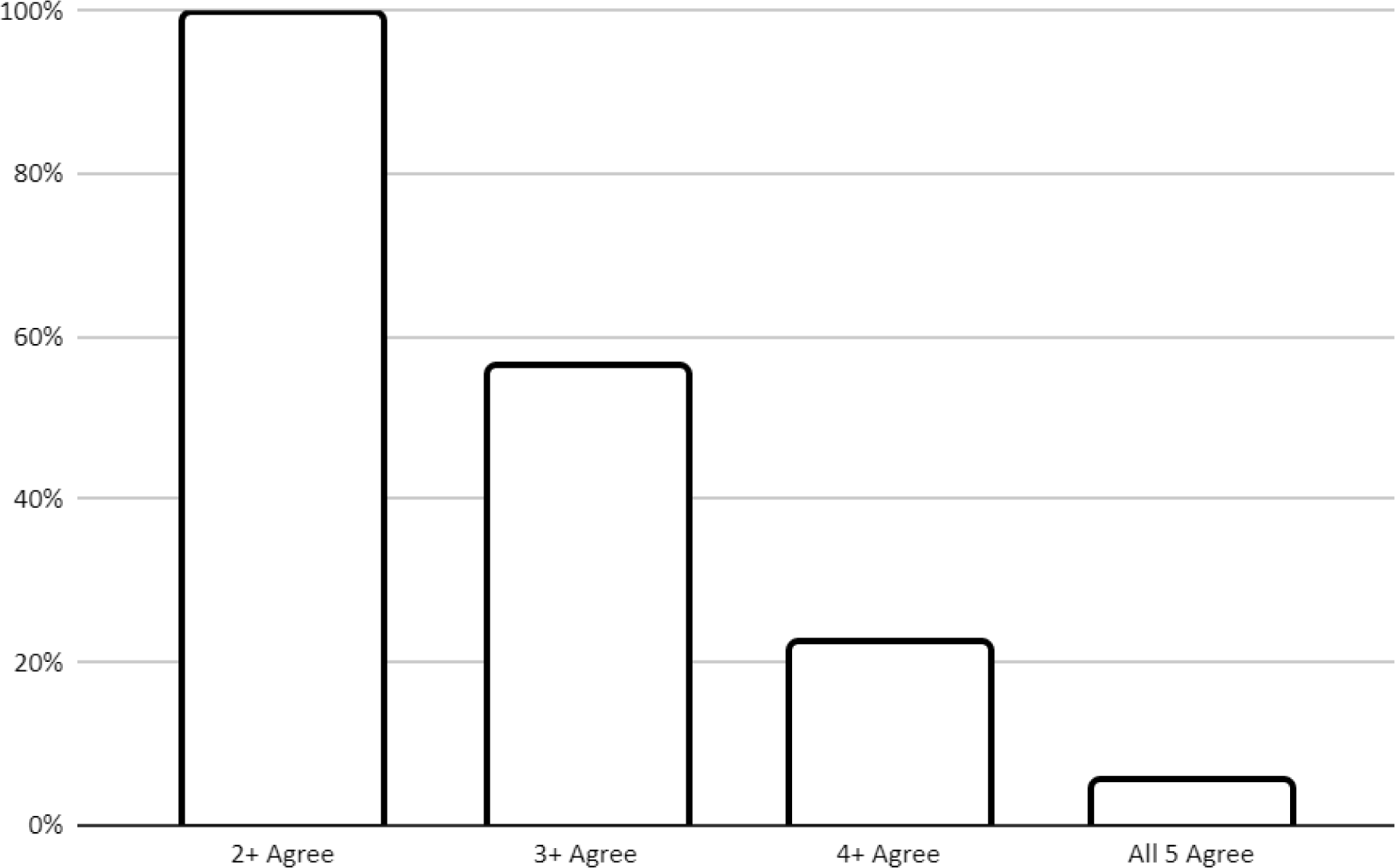
Model agreement of most likely diagnosis category

The recommended initial test to order in the emergency department closely followed the initial diagnosis rather than clinical thinking. For example, if the model gave a most likely diagnosis of gastroesophageal reflux disease, the models would frequently recommend endoscopy as the initial test to order in the emergency department.

### Gender and racial bias analysis

There was no gender or racial bias noted regarding the most likely diagnosis or initial test recommended. An EKG was recommended as the first test regardless of gender or race if the most likely diagnosis was cardiovascular. For example, for model five, an EKG was recommended as the initial test 64% of the time for men and 65% of the time for women; for African Americans, an EKG was recommended 65.0% of the time, and for non-African Americans, an EKG was also recommended 65.0% of the time.

There was evidence of possible bias in terms of the weights assigned to the variables of gender and race when determining a risk score. Being male and/or African American, on average, increased the risk of acute coronary syndrome by the five models built upon the third dataset consisting of the forty-four history and physical variables. By these weights, being male increased the risk score by 3.2% on average, and being African American increased the risk score by 1.3% on average.

## Discussion

This study found that ChatGPT-4 gives highly variable risk estimates, diagnostic categories, and recommendations for test ordering when presented with the exact same clinical data. This variation is so large that if applied to clinical practice, patient care would be unpredictable compared to well-validated scoring systems such as TIMI and HEART.

In head-to-head comparisons, the mean ChatGPT score was slightly but significantly higher than the TIMI score (3.55 vs 3.50, p < 0.001), although the correlation between the two was strong (r = 0.989, p < 0.001). However, more importantly, ChatGPT gave a different score from TIMI 45% of the time and widely varying scores when presented with identical risk data over five separate trials. Although the mean TIMI and ChatGPT scores are nearly identical, the wide distribution of ChatGPT scores compared to TIMI raises concerns about the current reliability of this model in predicting cardiac risk.

The mean HEART score was higher than the ChatGPT score (4.99 versus 4.92, p < 0.001). However, the distribution of scores given by ChatGPT was again broad, resulting in ChatGPT giving a different score from HEART 48% of the time. Furthermore, for low-risk ChatGPT scores, HEART gave a higher risk category 24% of the time, and for low-risk HEART scores, ChatGPT gave a higher risk category 12% of the time. Given that the primary clinical utility of HEART is to help identify low-risk patients suitable for an outpatient workup, this disagreement is concerning. If applied clinically, one out of four patients categorized as low risk by ChatGPT would be categorized as moderate or high risk by HEART.

Both TIMI and HEART rely on test results to risk stratify patients. So, to see how ChatGPT approached simulated patients at presentation, the AI was asked to analyze 44 history and physical variables related to acute nontraumatic chest pain. Again, ChatGPT gave significantly different responses when presented with the same data on multiple occasions. The complexity of 44 variables resulted in an even broader distribution of risk scoring compared to the TIMI and HEART models. Although statistically significant, the correlation of the individual models with the average model was only moderate (r = 0.60). When the scores were normalized to a 10-point scale, the individual models differed from the average model 76% of the time. This large disagreement again supports the hypothesis that ChatGPT risk scoring of nontraumatic acute chest pain is inconsistent and thus unreliable. ChatGPT does even worse in determining the most likely diagnostic category, with a majority of the five models agreeing only 56% of the time. Furthermore, the recommendations for working up patients were frequently illogical. For example, ChatGPT often recommended upper endoscopy as the first test to order if the model suspected a gastrointestinal diagnosis.

We were interested to see if ChatGPT-4 might inadvertently have incorporated either racial or gender bias when evaluating the complaint of acute nontraumatic chest pain. Although being African American and/or male were both considered by ChatGPT to be cardiac risk factors, the small weights given to each suggest minimal racial or gender bias. ChatGPT did not diagnose cardiovascular disease more or less often based on race or gender and did not recommend cardiovascular testing more or less often based on race or gender.

Although analyses of other medical conditions have identified possible racial biases (23), our analysis found the opposite. Bias by LLMs comes from the data it trained on. In the case of the emergency evaluation of acute nontraumatic chest pain, gender bias was first identified in the early 1990s (24), leading to greater awareness and mitigation of this bias in the cardiovascular and emergency medicine literature. Nevertheless, ChatGPT did give extra weight to male gender, raising the possibility of a persistent bias in the training data affecting ChatGPT’s behavior.

ChatGPT is known to have a randomization factor as part of its algorithm for it to create natural language text. This is done through its temperature parameter, with a higher temperature resulting in more creative and varied inputs and a lower temperature producing more focused responses. The default temperature for ChatGPT’s web interface is 1.0, and this cannot currently be modified through the web interface. However, it can be changed via the application programming interface (API). While this randomness of

ChatGPT is well known regarding its generation of language, our study found that this randomness also affects its data analysis. To get more consistent and clinically reliable results, a customized program that accesses ChatGPT via API and sets the temperature to zero would need to be evaluated.

Accessing ChatGPT via API to lower its temperature, thus decreasing the randomness of output, would likely result in more consistent responses. However, the wide distribution of responses to the third dataset containing 44 history and physical variables suggests that ChatGPT needs to be trained more consistently. As an LLM, ChatGPT has been trained on the equivalent of hundreds of millions of books. While this gives ChatGPT a broad knowledge base, the large amount of training data also introduces contradictory, conflicting information. The latest iteration of ChatGPT, as of the time of this writing, introduces the ability to create specialized models trained on a limited set of highly curated data. For example, a specialized GPT model could be trained exclusively on PubMed Central articles or exclusively on recognized textbooks of emergency medicine, such as Rosen’s Emergency Medicine (25). This would decrease the effect of garbage in, garbage out (26) and could potentially revolutionize its clinical applications.

## Conclusion

Cardiovascular risk estimates by ChatGPT-4 on large, simulated patient datasets correlate well with the well-validated TIMI and HEART scores. Still, the variability of individual scores on identical risk data would make using uncustomized ChatGPT-4 out-of-the-box for cardiac risk assessment problematic. Setting the randomness (temperature) of the system to a lower setting and incorporating focused, curated training may result in risk assessment superior to currently used protocols for patients with acute nontraumatic chest pain.

